# Low daytime light and bright night-time light are associated with psychiatric disorders: an objective light study in >85,000 UK Biobank participants

**DOI:** 10.1101/2022.10.16.22280934

**Authors:** Angus C. Burns, Daniel P. Windred, Martin K. Rutter, Patrick Olivier, Céline Vetter, Richa Saxena, Jacqueline M. Lane, Andrew J. K. Phillips, Sean W. Cain

## Abstract

Circadian rhythm disturbance is a common feature of many psychiatric disorders. Light is the primary input to the circadian clock, with daytime light strengthening rhythms and night light disrupting them. Therefore, habitual light exposure may represent an environmental risk factor for susceptibility to psychiatric disorders. We performed the largest to-date cross-sectional analysis of light, sleep, physical activity, and mental health (*n* = 86,772 adults; aged 62.4 ± 7.4 years; 57% women). We examined the independent association of day and night light exposure with covariate-adjusted risk for psychiatric disorders and self-harm. Greater night light exposure was associated with increased risk for major depressive disorder, generalized anxiety disorder, PTSD, psychosis, bipolar disorder, and self-harm behavior. Independent of night light, greater day light exposure was associated with reduced risk for major depressive disorder, PTSD, psychosis, and self-harm behavior. These findings were robust to adjustment for sociodemographics, photoperiod, physical activity, and sleep quality. Avoiding light at night and seeking light during the day may be a simple and effective, non-pharmacological means of broadly improving mental health.

## Introduction

Healthy circadian rhythms are essential for mental health and wellbeing^1^. Many psychiatric disorders are characterized by disrupted circadian rhythms and sleep^2,3^. In humans, a central circadian (∼24-hour) clock in the suprachiasmatic nuclei (SCN) of the hypothalamus regulates the timing of basic cellular functions^4^, physiology, cognition, and behavior^5,6^. Rhythms within the SCN are regulated by daily light exposure patterns. This biological system evolved under predictable conditions of bright light during the day and darkness at night to ensure stable, robust rhythms^7,8^. Humans in modern, industrialized societies challenge this biology, spending ∼90% of the day indoors under electric lighting^9^ which is dim during the day and bright at night compared to natural light/dark cycles^10^. Deviations from our natural light/dark cycle lead to disrupted circadian rhythms and therefore could contribute to adverse psychiatric outcomes.

Here, we report the largest sample to-date (*n* = 86,772) of objective recordings of individual, 24-hour light exposure data. We investigated the association of day and night light with a constellation of psychiatric disorders that feature circadian rhythm disturbance, while controlling for confounding variables in multivariable models. We tested two primary hypotheses: (i) greater light exposure in the day is associated with lower risk for psychiatric disorders and better mood; and (ii) greater light exposure at night is associated with higher risk for psychiatric disorders and poorer mood. These hypotheses were motivated by the known effects of day and night light exposure on the human circadian system and the well-established links between circadian disruption and psychiatric disorders.

## Results

Of the 103,720 UK Biobank participants that completed the actigraphy assessment, we excluded those with poor quality or unreliable accelerometry, sleep, and light data (detailed in Supplementary Methods and Supplementary Figure 1), leaving 86,772 participants. Of these, 86,631 had complete data for day and night light as well as the covariates in our fully adjusted Model 3. Characteristics of the sample and their missingness are presented in Table 1 for the lowest (Q1 & Q2) and highest (Q3 & Q4) quartiles of day and night light exposure. The light measurements showed good reliability, intra-class correlation (ICC) = 0.82 (95% CI 0.81-0.83; see Supplementary Methods). The analytical sample comprised participants who completed both the MHQ and actigraphy assessment and sample sizes varied depending on the outcome variable assessed, ranging from 26,956 to 61,442 for Model 1 and 26,824 to 61,147 in Model 3. Summary statistics for outcome variables derived from the MHQ are detailed in Supplementary Tables 1 and 2.

**Table 1.** Demographic characteristics of participants by low and high day and night light exposure

|  | Day light |  |  | Night light |  |  |
| --- | --- | --- | --- | --- | --- | --- |
|  | Low<br>Q1 & Q2<br>N=43386 | High<br>Q3 & Q4<br>N=43386 | *p*<br>value | Low<br>Q1 & Q2<br>N=43387 | High<br>Q3 & Q4<br>N=43385 | *p*<br>value |
| **Age** |  |  |  |  |  |  |
| Mean (SD) | 62.0 (7.94) | 62.7 (7.71) | <0.0001 | 62.6 (7.86) | 62.1 (7.79) | <0.0001 |
| Median [IQR] | 62.9 [55.7, 68.4] | 64.0 [57.0, 68.7] |  | 63.8 [56.6, 68.8] | 63.2 [56.1, 68.3] |  |
| **Sex** |  |  |  |  |  |  |
| Female | 25159 (58.0%) | 24321 (56.1%) | <0.0001 | 25352 (58.4%) | 24128 (55.6%) | <0.0001 |
| Male | 18227 (42.0%) | 19065 (43.9%) |  | 18035 (41.6%) | 19257 (44.4%) |  |
| **BMI** |  |  |  |  |  |  |
| Mean (SD) | 26.7 (4.57) | 26.7 (4.47) | 0.21 | 26.3 (4.31) | 27.1 (4.68) | <0.0001 |
| Median [IQR] | 26.0 [23.6, 29.1] | 26.1 [23.6, 29.0] |  | 25.7 [23.3, 28.6] | 26.4 [23.9, 29.5] |  |
| Missing | 102 (0.2%) | 83 (0.2%) |  | 83 (0.2%) | 102 (0.2%) |  |
| **White ethnicity** |  |  |  |  |  |  |
|  | 41679 (96.1%) | 42226 (97.3%) | <0.0001 | 42233 (97.3%) | 41672 (96.1%) | <0.0001 |
| Missing | 150 (0.3%) | 141 (0.3%) |  | 136 (0.3%) | 155 (0.4%) |  |
| **Employment** |  |  |  |  |  |  |
| No | 15912 (36.7%) | 17412 (40.1%) | <0.0001 | 17353 (40.0%) | 15971 (36.8%) | <0.0001 |
| Yes | 27474 (63.3%) | 25974 (59.9%) |  | 26034 (60.0%) | 27414 (63.2%) |  |
| **Shiftwork** |  |  |  |  |  |  |
| No | 39780 (91.7%) | 40152 (92.5%) | <0.0001 | 40545 (93.4%) | 39387 (90.8%) | <0.0001 |
| Yes | 3606 (8.3%) | 3234 (7.5%) |  | 2842 (6.6%) | 3998 (9.2%) |  |
| **Physical activity** |  |  |  |  |  |  |
| Mean (SD) | 27.3 (7.89) | 29.0 (8.16) | <0.0001 | 28.1 (7.97) | 28.3 (8.17) | 0.0003 |
| Median [IQR] | 26.4 [21.9, 31.7] | 28.1 [23.4, 33.6] |  | 27.1 [22.6, 32.5] | 27.3 [22.6, 32.8] |  |
| Missing | 39 (0.1%) | 94 (0.2%) |  | 60 (0.1%) | 73 (0.2%) |  |
| **Photoperiod** |  |  |  |  |  |  |
| Mean (SD) | 10.9 (2.87) | 14.1 (2.58) | <0.0001 | 12.4 (3.06) | 12.6 (3.28) | <0.0001 |
| Median [IQR] | 10.2 [8.47, 12.8] | 14.8 [12.3, 16.4] |  | 12.4 [9.58, 15.2] | 12.8 [9.61, 15.9] |  |
| **Sleep duration** |  |  |  |  |  |  |
| Mean (SD) | 7.56 (1.06) | 7.58 (1.02) | 0.004 | 7.79 (0.95) | 7.34 (1.07) | <0.0001 |
| Median [IQR] | 7.60 [6.95, 8.21] | 7.61 [6.98, 8.22] |  | 7.81 [7.23, 8.38] | 7.37 [6.72, 8.00] |  |
| **Sleep efficiency** |  |  |  |  |  |  |
| Mean (SD) | 0.88 (0.06) | 0.88 (0.06) | <0.0001 | 0.89 (0.06) | 0.88 (0.07) | <0.0001 |
| Median [IQR] | 0.89 [0.85, 0.92] | 0.90 [0.86, 0.92] |  | 0.90 [0.86, 0.93] | 0.89 [0.85, 0.92] |  |
| **Urban residence** |  |  |  |  |  |  |
| Yes | 36703 (84.6%) | 35533 (81.9%) | <0.0001 | 35687 (82.3%) | 36549 (84.2%) | <0.0001 |
| Missing | 430 (1.0%) | 404 (0.9%) |  | 423 (1.0%) | 411 (0.9%) |  |

IQR = interquartile range, SD = standard deviation. *p* values correspond to *t* tests for continuous variables and $\chi^2$ tests for categorical variables.

In fully adjusted regression models (Model 3, adjusted for age, sex, ethnicity, photoperiod, employment, and physical activity; Figure 1 & Table 2), higher night light exposure was associated with higher odds of major depressive disorder (χ^2^ *P* < 0.0001; fourth quartile odds ratio (OR_Q4_) = 1.30, 95% CI = 1.23-1.38), self-harm (χ^2^ *P* < 0.0001; OR_Q4_ = 1.27, 95% CI = 1.14-1.42), generalized anxiety disorder (χ^2^ *P* = 0.0001; OR_Q4_ = 1.23, 95% CI = 1.11-1.36), PTSD (χ^2^ *P* < 0.0001; OR_Q4_ = 1.34, 95% CI = 1.22-1.48), and psychosis (χ^2^ *P* = 0.0009; OR_Q4_ = 1.21, 95% CI = 1.09-1.34). There was no overall association with bipolar disorder (χ^2^ *P* = 0.18), however those in the brightest night light quartile had 1.20 (95% CI = 1.02-1.42) times higher risk. Greater night light exposure was associated with higher scores on the PHQ-9 (χ^2^ *P* < 0.0001; Standardized β_Q4_ = 0.13, 95% CI = 0.11-0.15), GAD-7 (χ^2^ *P* < 0.0001; β_Q4_ = 0.07, 95% CI = 0.05-0.09), PCL-6 (χ^2^ *P* < 0.0001; β_Q4_ = 0.11, 95% CI = 0.08-0.14), and lower wellbeing scores (χ^2^ *P* < 0.0001; β_Q4_ = −0.11, 95% CI = −0.13--0.08; Figure 2, Supplementary Table 3). The results for Model 1 (unadjusted) and Model 2 (adjusted for age, sex, ethnicity, and photoperiod) were similar in terms of direction, strength, and significance to Model 3.

**Table 2.** Associations between night light exposure and psychiatric outcomes.

| Night light | Model 1<br>OR (95% CI) |  |  |  | Model 2<br>aOR (95% CI)* |  |  |  | Model 3<br>aOR (95% CI)<sup>†</sup> |  |  |  |
| --- | --- | --- | --- | --- | --- | --- | --- | --- | --- | --- | --- | --- |
| | 2<sup>nd</sup><br>Quartile | 3<sup>rd</sup><br>Quartile | 4<sup>th</sup><br>Quartile | LRT $p^{\ddagger}$ | 2<sup>nd</sup><br>Quartile | 3<sup>rd</sup><br>Quartile | 4<sup>th</sup><br>Quartile | LRT $p^{\ddagger}$ | 2<sup>nd</sup><br>Quartile | 3<sup>rd</sup><br>Quartile | 4<sup>th</sup><br>Quartile | LRT $p^{\ddagger}$ |
| Major depressive disorder | 1.05<br>(1.00 – 1.11) | **1.17**<br>(**1.11 – 1.24**) | **1.28**<br>(**1.22 – 1.36**) | **<0.0001** | 1.04<br>(0.98 – 1.09) | **1.17**<br>(**1.11 – 1.24**) | **1.32**<br>(**1.25 – 1.39**) | **<0.0001** | 1.04<br>(0.98 – 1.10) | **1.18**<br>(**1.12 – 1.25**) | **1.30**<br>(**1.23 – 1.38**) | **<0.0001** |
| Self-harm | 1.02<br>(0.91 – 1.14) | 1.03<br>(0.92 – 1.15) | **1.28**<br>(**1.15 – 1.42**) | **<0.0001** | 0.99<br>(0.88 – 1.10) | 0.99<br>(0.88 – 1.11) | **1.28**<br>(**1.15 – 1.43**) | **<0.0001** | 0.99<br>(0.88 – 1.11) | 1.00<br>(0.89 – 1.12) | **1.27**<br>(**1.14 – 1.42**) | **<0.0001** |
| Generalized anxiety disorder | 1.10<br>(0.99 – 1.21) | **1.22**<br>(**1.10 – 1.35**) | **1.23**<br>(**1.11 – 1.36**) | **<0.0001** | 1.06<br>(0.96 – 1.18) | **1.18**<br>(**1.07 – 1.31**) | **1.23**<br>(**1.11 – 1.36**) | **0.0002** | 1.07<br>(0.97 – 1.19) | **1.20**<br>(**1.08 – 1.33**) | **1.23**<br>(**1.11 – 1.36**) | **0.0001** |
| PTSD | 1.03<br>(0.93 – 1.13) | **1.19**<br>(**1.08 – 1.31**) | **1.35**<br>(**1.23 – 1.48**) | **<0.0001** | 1.00<br>(0.90 – 1.11) | **1.15**<br>(**1.04 – 1.27**) | **1.35**<br>(**1.22 – 1.48**) | **<0.0001** | 1.00<br>(0.91 – 1.11) | **1.16**<br>(**1.05 – 1.29**) | **1.34**<br>(**1.22 – 1.48**) | **<0.0001** |
| Bipolar disorder | 1.15<br>(0.98 – 1.36) | 1.17<br>(0.99 – 1.38) | **1.25**<br>(**1.06 – 1.47**) | 0.06 | 1.11<br>(0.94 – 1.31) | 1.11<br>(0.94 – 1.31) | **1.21**<br>(**1.02 – 1.43**) | 0.18 | 1.11<br>(0.94 – 1.31) | 1.12<br>(0.95 – 1.32) | **1.20**<br>(**1.02 – 1.42**) | 0.18 |
| Psychosis | 1.05<br>(0.95 – 1.17) | **1.17**<br>(**1.05 – 1.29**) | **1.21**<br>(**1.10 – 1.35**) | **0.0005** | 1.03<br>(0.93 – 1.15) | **1.13**<br>(**1.02 – 1.26**) | **1.22**<br>(**1.10 – 1.35**) | **0.0004** | 1.04<br>(0.93 – 1.15) | **1.14**<br>(**1.03 – 1.27**) | **1.21**<br>(**1.09 – 1.34**) | **0.0009** |

Note: logistic regression was used for all outcomes and odds ratios (ORs) are presented with their 95% confidence intervals for each ascending night light quartile relative to the low light (Q1) referent. Model 1 is unadjusted. \*Model 2 adjusted for age, sex, ethnicity, and photoperiod as a measure of seasonality. <sup>†</sup>Model 3 is additionally adjusted for employment and physical activity. Bolded coefficients indicate significance at the .05 level.

**Figure 1.**
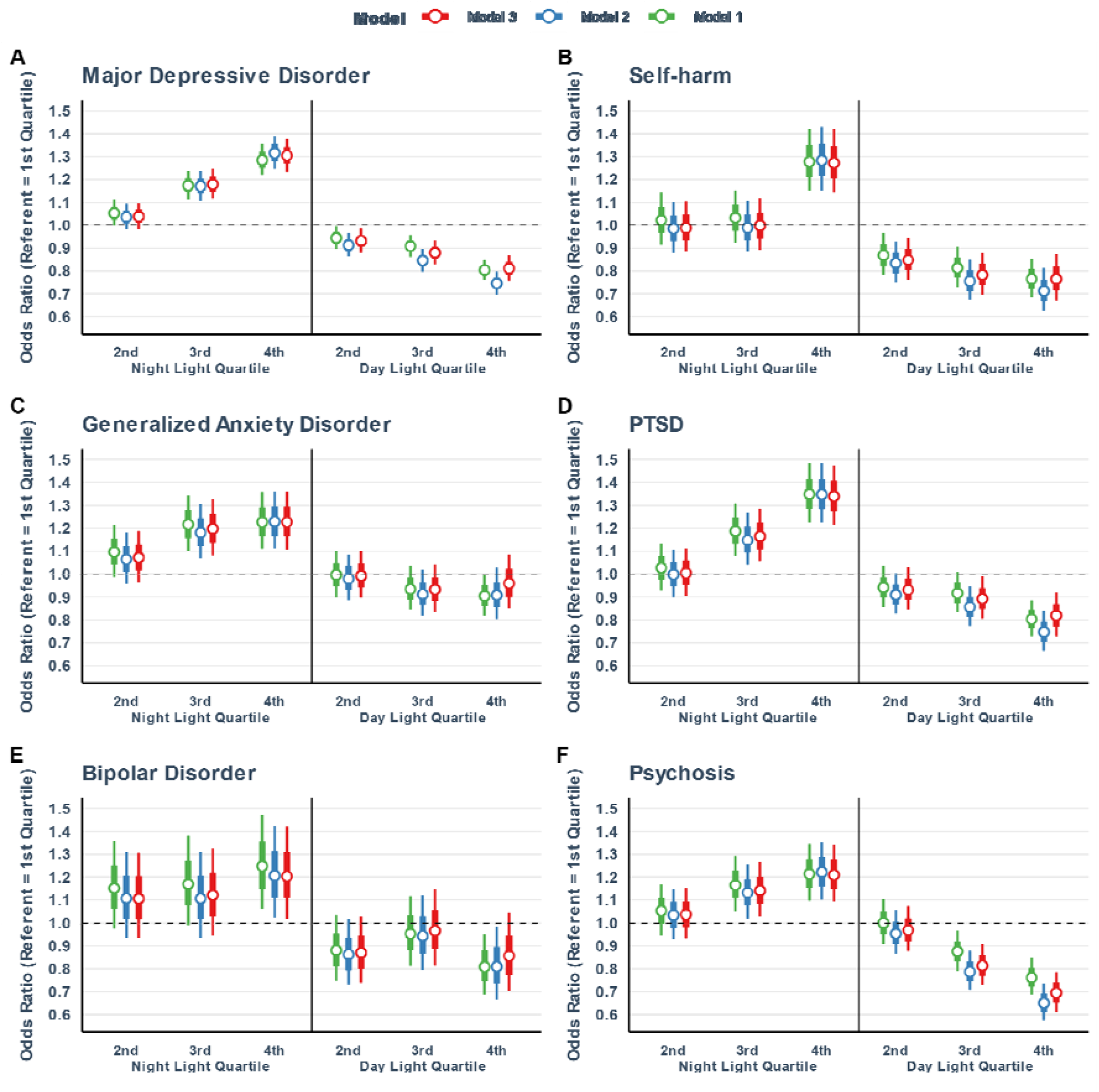
Associations of day and night light exposure with psychiatric disorders and self-harm. Coefficients plot for day and night-time light associations with (A) major depressive disorder, (B) self-harm behavior, (C) generalized anxiety disorder, (D) PTSD, (E) bipolar disorder and (F) psychosis. Coefficients represent the odds ratios ± the SEM (inner error bar) and 95% CI (outer error bar) for each quartile of day and night light exposure relative to the low light (Q1) referent. Three models are presented with increasing adjustment for confounders: Model 1 (green) is unadjusted, Model 2 (blue) adjusts for age, sex, ethnicity and photoperiod and Model 3 (red) additionally adjusts for employment and physical activity.

**Figure 2.**
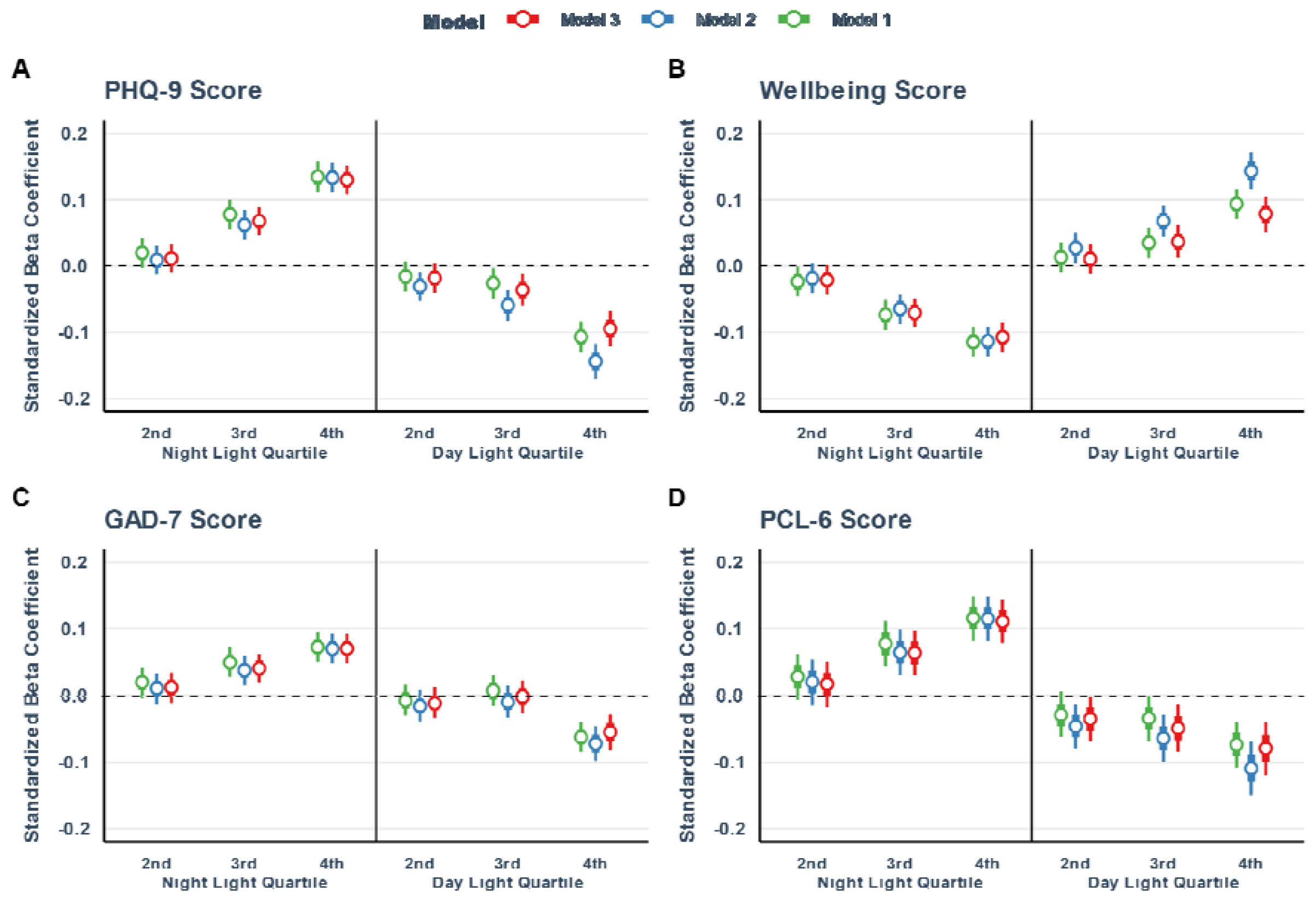
Associations of day and night light exposure with symptom severity scales and wellbeing. Coefficients plot for day and night-time light associations with (A) PHQ-9 score, (B) wellbeing score, (C) GAD-7 score and (D) PCL-6 score. Coefficients represent the standardized betas ± the SEM (inner error bar) and 95% CI (outer error bar) for each quartile of day and night light exposure relative to the low light (Q1) referent. Three models are presented with increasing adjustment for confounders: Model 1 (green) is unadjusted, Model 2 (blue) adjusts for age, sex, ethnicity and photoperiod and Model 3 (red) additionally adjusts for employment and physical activity.

Higher day light exposure (Model 3; Figure 1 & Table 3), was associated with lower odds of major depressive disorder (χ^2^ *P* < 0.0001; OR_Q4_ = 0.81, 95% CI = 0.76-0.87), self-harm (χ^2^ *P* = 0.0001; OR_Q4_ = 0.76, 95% CI = 0.67-0.87), PTSD (χ^2^ *P* = 0.01; OR_Q4_ = 0.82, 95% CI = 0.73-0.92), and psychosis (χ^2^ *P* < 0.0001; OR_Q4_ = 0.69, 95% CI = 0.61-0.79). There was no association of day light exposure with generalized anxiety disorder (χ^2^ *P* = 0.61; OR_Q4_ = 0.96, 95% CI = 0.85-1.09) or lifetime bipolar disorder (χ^2^ *P* = 0.22; OR_Q4_ = 0.86, 95% CI = 0.70-1.05). Figure 2 and Supplementary Table 4 show that greater day light exposure was associated with lower scores on the PHQ-9 (χ^2^ *P* < 0.0001; β_Q4_ = −0.09, 95% CI = −0.12--0.07), GAD-7 (χ^2^ *P* < 0.0001; β_Q4_ = −0.05, 95% CI = −0.08--0.03), PCL-6 (χ^2^ *P* = 0.002; β_Q4_ = −0.08, 95% CI = −0.12--0.04), and higher wellbeing scores (χ^2^ *P* < 0.0001; β_Q4_ = 0.08, 95% CI = 0.05-0.11). The results for Model 1 (unadjusted) and Model 2 (adjusted for age, sex, ethnicity, and photoperiod) were similar in terms of direction and significance to Model 3. However, ORs and betas tended to be smaller in Model 3 after the addition of physical activity and employment covariates.

**Table 3.** Associations between day light exposure and psychiatric outcomes.

|  | Model 1<br>OR (95% CI) |  |  |  | Model 2<br>aOR (95% CI)* |  |  |  | Model 3<br>aOR (95% CI)<sup>†</sup> |  |  |  |
| --- | --- | --- | --- | --- | --- | --- | --- | --- | --- | --- | --- | --- |
| Day light | 2<sup>nd</sup><br>Quartile | 3<sup>rd</sup><br>Quartile | 4<sup>th</sup><br>Quartile | LRT *p*<sup>‡</sup> | 2<sup>nd</sup><br>Quartile | 3<sup>rd</sup><br>Quartile | 4<sup>th</sup><br>Quartile | LRT *p*<sup>‡</sup> | 2<sup>nd</sup><br>Quartile | 3<sup>rd</sup><br>Quartile | 4<sup>th</sup><br>Quartile | LRT *p*<sup>‡</sup> |
| Major depressive disorder | **0.94**<br>(0.90 – 1.00) | **0.91**<br>(0.86 – 0.96) | **0.80**<br>(0.76 – 0.85) | <0.0001 | **0.91**<br>(0.86 – 0.97) | **0.85**<br>(0.80 – 0.90) | **0.75**<br>(0.70 – 0.80) | <0.0001 | **0.93**<br>(0.88 – 0.99) | **0.88**<br>(0.83 – 0.93) | **0.81**<br>(0.76 – 0.87) | <0.0001 |
| Self-harm | **0.87**<br>(0.78 – 0.97) | **0.81**<br>(0.73 – 0.90) | **0.77**<br>(0.69 – 0.85) | <0.0001 | **0.83**<br>(0.75 – 0.93) | **0.76**<br>(0.67 – 0.85) | **0.71**<br>(0.63 – 0.81) | <0.0001 | **0.85**<br>(0.76 – 0.94) | **0.78**<br>(0.70 – 0.88) | **0.76**<br>(0.67 – 0.87) | 0.0001 |
| Generalized anxiety disorder | 1.00<br>(0.90 – 1.10) | 0.94<br>(0.85 – 1.03) | 0.91<br>(0.82 – 1.00) | 0.14 | 0.98<br>(0.89 – 1.09) | 0.91<br>(0.82 – 1.02) | 0.91<br>(0.81 – 1.03) | 0.28 | 0.99<br>(0.90 – 1.10) | 0.93<br>(0.84 – 1.04) | 0.96<br>(0.85 – 1.09) | 0.61 |
| PTSD | 0.94<br>(0.86 – 1.04) | 0.92<br>(0.83 – 1.01) | **0.80**<br>(0.73 – 0.89) | **0.0001** | 0.91<br>(0.83 – 1.00) | **0.86**<br>(0.77 – 0.95) | **0.75**<br>(0.67 – 0.84) | <0.0001 | 0.93<br>(0.85 – 1.03) | **0.89**<br>(0.81 – 0.99) | **0.82**<br>(0.73 – 0.92) | **0.01** |
| Bipolar disorder | 0.88<br>(0.75 – 1.04) | 0.95<br>(0.81 – 1.12) | **0.81**<br>(0.69 – 0.95) | 0.06 | 0.86<br>(0.73 – 1.02) | 0.94<br>(0.80 – 1.12) | **0.81**<br>(0.67 – 0.99) | 0.10 | 0.87<br>(0.74 – 1.03) | 0.97<br>(0.81 – 1.15) | 0.86<br>(0.70 – 1.05) | 0.22 |
| Psychosis | 1.00<br>(0.91 – 1.10) | **0.88**<br>(0.79 – 0.97) | **0.76**<br>(0.69 – 0.85) | <0.0001 | 0.95<br>(0.86 – 1.06) | **0.79**<br>(0.71 – 0.88) | **0.65**<br>(0.57 – 0.74) | <0.0001 | 0.97<br>(0.88 – 1.07) | **0.81**<br>(0.73 – 0.91) | **0.69**<br>(0.61 – 0.79) | <0.0001 |

Note: logistic regression was used for all outcomes and odds ratios (ORs) are presented with their 95% confidence intervals for each ascending day light quartile relative to the low light (Q1) referent. Model 1 is unadjusted. \*Model 2 adjusted for age, sex, ethnicity, and photoperiod as a measure of seasonality. <sup>†</sup>Model 3 is additionally adjusted for employment and physical activity. Bolded coefficients indicate significance at the .05 level.

A series of sensitivity analyses were completed. First, we re-ran Model 3 regressions excluding participants who reported doing shift work (*n* = 6,840, 7.9%). The results for both psychiatric disorders and symptom severity scores were unchanged in this subsample (Supplementary Tables 5 and 6). The second sensitivity analysis examined whether observed associations of day and night light with symptom severity scales and wellbeing were driven by the presence of clinical subgroups. We re-ran these analyses excluding participants with relevant disorders (see Methods), adjusting for the same covariates in Model 3. The associations were unchanged, excepting small variations in strength (Supplementary Table 7). The third sensitivity analysis examined whether the associations between light and psychiatric outcomes were independent of sleep characteristics. After adjustment for actigraphically derived sleep duration and sleep efficiency in addition to the Model 3 covariates (Model 4) the association of night light exposure with psychosis was no longer significant (χ^2^ *P* < 0.09; OR_Q4_ = 1.12, 95% CI = 1.01-1.25). All other associations of day and night light with psychiatric outcomes and symptom severity scales were unchanged, though associations tended to be weaker (Supplementary Tables 8 and 9). Finally, to assess whether residential density as a measure of urbanicity could explain the light and mood relationship, we conducted an additional sensitivity analysis (Model 5) adjusting for residence type (urban vs. rural) as well as the covariates in Model 3. Adjustment for urbanicity did not change any of the associations of day and night light with psychiatric outcomes and symptom severity scales in strength or significance (Supplementary Tables 10 and 11).

## Discussion

We found that objectively measured light exposure patterns under free-living conditions were associated with the risk for psychiatric disorders and the severity of mood symptoms. Brighter light at night was associated with a greater risk for major depressive disorder, self-harm behavior, PTSD, psychosis, generalized anxiety disorder, and bipolar disorder, as well as poorer self-reported mood and wellbeing. Conversely, brighter light in the day was associated with lower odds of major depressive disorder, self-harm behavior, PTSD, and psychosis, as well as better self-reported mood and wellbeing. Remarkably, these associations were independent and additive. For example, greater night light exposure was associated with increased odds of major depressive disorder even for those in the brightest day light quartile. These associations were also independent of demographic, physical activity, photoperiod, and employment covariates. Sensitivity analyses showed these findings to be consistent when accounting for shift work, sleep, and urbanicity.

For individuals in the brightest night light quartile, we observed ∼30% higher risk of MDD and self-harm, while individuals in the brightest day light quartile had ∼20% lower risk of MDD and self-harm. Night light exposure was also associated with poorer self-reported mood and wellbeing, while day light exposure was associated with better mood and wellbeing. Previous studies have reported an association of night light exposure with low mood^11^ and one study has linked night light to MDD risk, though this study examined group-level, outdoor light at night which may not be a good proxy for individual, ambient light levels^12^. Other limitations of these studies were generally small sample sizes, poor control for confounders such as physical activity and sleep quality, and no studies considered the independent effects of day and night light. Conversely, day light therapy has long been shown to be efficacious in treating depression^13^ and has been shown to significantly enhance treatment efficacy when combined with an SSRI^14^. Fewer studies have linked free-living day light exposure to MDD risk, though one study linked self-reported time spent in outdoor light with lower risk^15^. Depression has long been associated with circadian disruption. Patients with depression have both delayed and low amplitude circadian rhythms^16,17^ which are reversed in recovery^18^. The severity of mood symptoms and the duration of depressive episodes are greater in those who experience circadian rhythm disturbance^16,19^ and the presence of circadian rhythm disturbance in depression is predictive of recurrence^20^. Our findings are consistent with the known time-dependent effects of light on the properties of the circadian system, such that light at night tends to delay rhythms and reduce circadian amplitude, whereas early morning and day light tends to advance rhythms and boost circadian amplitude^8,21,22^. Therefore, the euthymic effect of bright day light and dim night light exposure may occur by boosting the amplitude and advancing the timing of circadian rhythms, correcting the delayed and blunted rhythms seen in depression^17,23^. Seeking greater day light and minimizing night light exposure could be a simple means of improving depression trajectories by treating underlying circadian disturbance.

In people with high levels of light exposure at night, we observed ∼20% higher risk for bipolar disorder. Bipolar disorder has long been associated with dampened amplitude of behavioral rhythms and more variable circadian timing^3,24^. A recent study found that brighter night light exposure predicted manic/hypomanic episodes in bipolar patients^25^ and outdoor light at night has been associated with bipolar disorder^12^. Hypersensitivity of the circadian system to light at night has been proposed to be a trait marker of bipolar disorder^26^. Drugs used to treat bipolar disorder reduce the sensitivity of the circadian system to light^27^, suggesting that reducing the effects of light at night on the circadian system may play a role in recovery. Consistent with this, nighttime dark therapy and wearing blue light blocking glasses at night are effective at reducing mania in patients^28,29^. We did not see an association of bipolar disorder with day light exposure. This finding is novel, as the association of day light with manic symptoms in bipolar disorder and the efficacy of day light therapy on manic episodes has not been examined^30^.The avoidance of light at night specifically may be beneficial in mitigating risk for bipolar disorder.

We found both an adverse association of night light exposure and a beneficial association of day light exposure with PTSD risk and symptom severity. To our knowledge, no studies have examined the association of free-living light exposure patterns in the day or night to PTSD risk. There is some evidence of disturbed circadian rhythms in PTSD. Delayed activity rhythms are associated with more severe PTSD^31^ symptoms while lower urinary melatonin rhythm amplitude after a trauma exposure predicts a higher risk for PTSD^32^, a finding replicated in military personnel^33^. Our results suggest bright night and dim day light exposure may be antecedent factors leading to blunted and delayed rhythms. This is supported by evidence that day light therapy may be an effective treatment for PTSD symptoms^34^. Avoidance of light at night and seeking bright day light after trauma could reduce the risk of developing PTSD or the severity of symptoms in those with the disorder.

Finally, bright night light exposure was associated with ∼20% increased risk for psychosis, while bright day light exposure was associated with ∼30% reduced risk for psychosis. There is also little data linking free-living light exposure in the day or night to psychosis and psychotic disorders, despite sleep and circadian rhythm disruption being common features of patients on and off medication^1^. One small study found that patients with schizophrenia had lower daytime light exposure and in a natural experiment observed that boosting daytime light exposure could normalize the sleep and circadian disruption seen in the disorder^35^. Studies of day light therapy and schizophrenia have reported mixed results, though these studies have been small and further, more rigorous trials are needed^36,37^. In addition to day light, our findings point to night light as a novel therapeutic target for psychosis.

Beyond effects on the circadian clock, non-visual photoreception is appreciated to have a direct effect on mood via projections to brain areas implicated in mood regulation. Light exposure acutely enhances both mood and alertness^38,39^. Intrinsically photosensitive retinal ganglion cells (ipRGCs) expressing the photopigment melanopsin are the primary input of light information to the circadian clock in the SCN^40^. These cells also project to the medial amygdala and lateral habenula, brain areas implicated in depression, and these projections mediate the acute euthymic effect of light exposure^41,42^. The direct effects of light may partially explain the association of day light with lower MDD and self-harm risk, though a mechanism for direct effects of day light on other disorders such as PTSD, psychosis, and bipolar disorder is unclear. As mood is generally poorer in the night/early morning hours, people may seek out the acute euthymic effects of light at night. Though this may immediately improve mood, it would lead to circadian disruption in the long term and could perpetuate mental illness. This represents a potential challenge for promoting healthy light behaviors.

Taken together, our findings are consistent with bright day light and low night light strengthening circadian rhythms as an antecedent to more robust mental health. Patterns of bright day light and low night light serve to enhance the amplitude and stability of the circadian clock as well as align its activity appropriately with daily activities^7,8,21^. As modern humans spend ∼90% of the day indoors^9^, our light exposure patterns are typically less bright in the day and more bright at night than naturalistic patterns across our evolutionary history^10^. Addressing this deviation from our natural light/dark cycles may improve the general mental health of people in industrialized societies.

This study has a number of important limitations. First, the findings we report are cross-sectional. While there are well-supported causal mechanisms linking bright night light and dim day light with circadian disruption, and linking circadian disruption with mental health, we acknowledge the possibility of reverse causation and that longitudinal studies will be needed to establish the temporality of the associations we observed. However, the robustness of our findings to adjustment for confounders including physical activity and sleep provides support for our interpretation. Second, light monitoring was performed using a wrist-worn device, which is not designed to measure light at the ocular level. The data therefore provide a coarse estimate of the actual effects of light on the circadian system. Third, the actigraphy data and outcome variables were not measured simultaneously, with the latter measured an average of 1.86 years later. We note that we did observe good reliability of within-individual repeat light exposure assessments completed over the course of a year, suggesting that light measurements at one timepoint are a good proxy for other timepoints.

This study is the first to report an analysis of objectively measured light exposure data from the UK Biobank and is the largest examination of objective light exposure and mental health to date. Our findings demonstrate a consistent association of healthy light exposure patterns with better psychiatric outcomes. These results suggest that brighter days and darker nights may be a simple, freely available, non-pharmacological intervention to enhance mental health.

## Supporting information

Supplementary Methods and Results

## Data Availability

This work utilized the UK Biobank resource (application 6818; Martin Rutter).

## Data availability statement

The data used in this study are available in the UK Biobank resource. Derived data supporting the findings of this study are available from the corresponding author (ACB) on request at.

## Methods

### Study design and participants

In this cross-sectional study we drew on the UK Biobank prospective general population cohort which contains more than 502,000 UK residents recruited via National Health Service (NHS) patient registers from 2006 to 2010. The study population is described in detail elsewhere^43,44^. Accelerometry and light data were measured in a subset of 103,720 participants in 2013-2015 and a separate subset of 157,366 participants completed an online Mental Health Questionnaire (MHQ) in 2016-2017. Participants who accepted the invitation to join the UK Biobank cohort provided written, informed consent and the UK Biobank has generic ethical approval from the North West Multi-Center Research Ethics Committee (ref 11/NW/03820).

### Measures

#### Light exposure

In 2013, 236,519 UK Biobank participants were invited to participate in a seven day physical activity and light monitoring study. Of these participants, 103,720 (43.9%) accepted, and returned the accelerometer to the UK Biobank. Participants who accepted the invitation received a wrist-worn AX3 triaxial accelerometer (Axivity, Newcastle upon Tyne, UK) with in-built light sensor (APDS9007 silicon photodiode sensor; spectral sensitivity λ = 470-650nm) and were asked to wear the device on their dominant wrist for seven days under free-living conditions.

Quality control and definition of light exposure predictors was completed with custom R (version 4.1.0) scripts detailed in the Supplementary Methods. Briefly, the daily light profiles of participants meeting quality control criteria were entered into a factor analysis to identify independent patterns in light exposure in the sample. Factor analysis supported the extraction of day (7.30am-8.30pm) and night (12.30am-6am) factors. Light exposure during the day had a small positive correlation with light exposure at night (*r*_*s*_ = 0.10, *p* < 0.0001). Due to large positive skew in both day and night light variables (skewness coefficient > 1 for both), both variables were converted into categorical predictors for analysis by dividing them into four equal-sized quartiles in ascending brightness.

#### Psychiatric outcomes

A total of 339,092 UK Biobank participants were invited to complete the online Mental Health Questionnaire (MHQ) in 2016 and 157,366 completed the questionnaire. Measurement of psychiatric outcomes as part of the MHQ took place an average of 1.86 years (SD = 0.66) after the actigraphy assessment. Of the 86,772 participants with complete light data, 61,466 (70.8%) completed the UK Biobank MHQ. Definition of case-control psychiatric disorder outcomes from the MHQ are based on Composite International Diagnostic Interview (CIDI) and Diagnostic and Statistical Manual IV (DSM-IV) criteria and followed guidelines established by Davis, Coleman ^45^. Case/control outcomes were major depressive disorder (MDD), generalized anxiety disorder (GAD), bipolar disorder, post-traumatic stress disorder (PTSD), psychosis, and self-harm. Detailed definitions are given in the Supplementary Methods. Continuous outcomes were symptom severity scales for depression (Patient Health Questionnaire, PHQ-9), anxiety (GAD-7), PTSD (PTSD Checklist-6, PCL-6), and an overall wellbeing score indexing euthymia and eudaemonia (see Supplementary Methods).

### Statistical analysis

The association between day and night light and case/control outcomes were examined with multiple logistic regression. Odds ratios (OR) and their 95% confidence intervals are reported. Multiple linear regression was used for the continuous symptom severity scales and wellbeing, with standardized beta values reported. These associations were tested hierarchically in three models with increasing adjustment for potential confounders. Each model included both day and night light categorical predictors to examine their independent effects. Likelihood-ratio χ^2^ tests were used as an omnibus test of significance for the day and night light factors. Model 1 examined the unadjusted association between day and night light and psychiatric outcomes. Model 2 adjusted for age (at the time of actigraphy), sex, ethnicity (white *vs*. non-white; data-field 21000), and photoperiod (as a measure of seasonality, defined as the duration between sunrise and sunset at the beginning of the actigraphy assessment). Finally, model 3 additionally adjusted for employment (employed *vs*. unemployed; data-field 6142) and physical activity (defined as overall acceleration average over the actigraphy period in milligravity units; data-field 90012). The physical activity variable was previously defined by the UK Biobank accelerometer expert working group^46^.

A series of sensitivity analyses were completed to examine the robustness of the primary findings. The first sensitivity analysis examined whether the presence of shift workers in the sample (*n* = 6,840, 7.9%) was driving the observed associations. Model 3 regressions were re-run excluding those who reported doing shiftwork (data-field 826) to any degree (sometimes, usually, or always). The second sensitivity analysis assessed whether observed associations of light with symptom severity scales and wellbeing were driven by clinical subgroups. We re-ran these analyses excluding participants with MDD for the PHQ-9 model; with GAD for the GAD-7 model; with PTSD for the PCL-6 model; and with MDD, GAD, PTSD, bipolar disorder, or psychosis for the wellbeing model. The third sensitivity analysis examined whether the relationship between light and psychiatric outcomes was independent of objectively measured sleep duration and efficiency (Model 4; see Supplementary Methods). The fourth sensitivity analysis examined whether participants’ residential population density as a measure of urbanicity could explain the light and mood relationship (Model 5). Urbanicity (data-field 20118; urban *vs*. rural) was defined according to the UK Office for National Statistics population density classification of participant residential postcodes where urban postcodes have a population of 10,000 or more and rural postcodes have a population of less than 10,000. Reporting of statistical analyses and results followed the STROBE guidelines.

