## Supplementary Methods and Results for "Low daytime light and bright night-time light are associated with psychiatric disorders: an objective light study in >85,000 UK Biobank participants"

**Supplementary Methods.** Accelerometry and sleep analysis, light analysis, and outcome variable definitions

**Supplementary Figure 1.** Flow diagram

**Supplementary Table 1.** Summary statistics of psychiatric outcomes among the light data sample

**Supplementary Table 2.** Summary statistics of symptom severity scales and wellbeing among the light data sample

**Supplementary Table 3.** Associations of night light exposure with symptom severity scales and wellbeing

**Supplementary Table 4.** Associations of day light exposure with symptom severity scales and wellbeing

**Supplementary Table 5.** Associations of day and night light exposure with psychiatric outcomes excluding shift workers

**Supplementary Table 6.** Associations of day and night light exposure with symptom severity scales and wellbeing excluding shift workers

**Supplementary Table 7.** Associations of day and night light exposure with symptom severity scales and wellbeing excluding participants meeting disorder criteria

**Supplementary Table 8.** Associations of day and night light exposure with psychiatric outcomes adjusting for sleep variables

**Supplementary Table 9.** Associations of day and night light exposure with symptom severity scales and wellbeing adjusting for sleep variables

**Supplementary Table 10.** Associations of day and night light exposure with psychiatric outcomes adjusting for urbanicity

**Supplementary Table 11.** Associations of day and night light exposure with symptom severity scales and wellbeing adjusting for urbanicity

**References.**

**Supplementary Methods.**

**Accelerometry and sleep analysis**

In 2013, 236,519 UK Biobank participants were invited to wear an accelerometer for 7 days as part of a physical activity and light monitoring study. Of these participants, 103,720 (43.9%) accepted, and returned the accelerometer to UK Biobank. Participants who accepted the invitation received a wrist-worn AX3 triaxial accelerometer (Axivity, Newcastle upon Tyne, UK) with in-built light sensor (APDS9007 silicon photodiode sensor; spectral sensitivity 𝜆 = 470-650nm) and were asked to wear the device on their dominant wrist for seven days under free-living conditions.

The R package GGIR^1,2^ (v1.6-9) was used to assess analyse accelerometry data, assess data quality and provide summaries of device non-wear and sleep parameters.

1. The raw accelerometry data files for each individual were downloaded in Continuous Wave Accelerometer (cwa) format and converted to Waveform Audio File (wav) format using the open-source software OMConvert.
2. Periods of device non-wear were identified algorithmically by GGIR^3^ and excluded from analysis
3. Sleep periods were then determined by GGIR using a validated, heuristic algorithm^1^, briefly:
a) The median z-angle (perpendicular to the wrist) was calculated in 5 second epochs using the three perpendicular axes of motion in relation to the downward force of gravity
b) A 5-minute rolling median of the absolute differences in z-angle between the 5 second epochs was calculated
c) The 10th percentile of the rolling median across a day (from noon to noon) was calculated and multiplied by 15 to set a threshold for determining inactivity
d) Blocks of inactivity were those for which the rolling median z-angle difference (from step b) is less than the threshold (from step c) and which last for 30 or more minutes
e) Blocks of inactivity (from step d) less than 60 minutes apart were treated as the same inactivity block
f) Labelling the longest combined period of sustained inactivity (from step e) in each day (noon-noon) as the sleep period time windows (SPT window) and all other periods of sustained inactivity as diurnal (daytime) inactivity. Note: if the SPT ends after noon (indicating sleep offset after noon) the sleep analysis is repeated on a 6am-6pm window in order to detect daytime sleepers.
g) Participants without any valid days of sleep-wake data due to file corruption or consistent non-wear interfering with SPT window detection were excluded (*n* = 8,055)
h) Sleep duration was calculated as the duration of GGIR-determined sustained inactivity between sleep onset and offset times. Where, sustained inactivity was defined as less than 5 degrees of movement from the accelerometer z-axis across rolling 5-minute intervals^4^.
i) Sleep efficiency was calculated as the proportion of sustained inactivity between sleep onset and offset, i.e. [total sustained inactivity between sleep onset and offset / (sleep offset time - sleep onset time)].

**Light analysis**

Following GGIR analysis, participants light data was mapped to the GGIR output and analysed with custom R scripts. Participants lacking any valid days of sleep-wake data due to data file corruption or consistent non-wear identified by GGIR were also excluded from light analysis (as noted above, *n* = 8,055).

1. Participants’ light data were extracted from the cwa files
2. Output current was then converted from the logarithmic scale to approximate lux according to the device manual (lux = 10^(device output current/341)
3. Output current of the light sensor was down-sampled from 100Hz to 1Hz and then averaged into 10-second epochs
4. Epochs of device non-wear identified by GGIR were marked as missing data
5. Participants with consistently dark (percentage of data below 1lux > 80%) or consistently bright (percentage of data above 600lux >75%) were excluded (*n* = 7,651), indicating device malfunction or coverage
6. Daily light profiles of participants were constructed by averaging epochs into forty-eight 30-min bins across the 24-hour period per study day and these bins were then averaged across the study days to generate an average 24-hour light exposure profile per-participant (i.e. each 30-minute bin contained the average light exposure at that time across the available study days).
7. Participants with less than two days’ worth of light data per 30-minute bin were excluded (*n* = 1,242) for a final sample of 86,772

Light exposure predictors were defined by factor analysis of the forty-eight 30-minute light bins across the 24-hour period. Factor analysis supported the extraction of a two-factor structure (day, 7.30am-8.30pm; and night, 12.30am-6am; factor loadings ≥ 0.5, varimax rotation, cumulative proportion of variance explained = 0.56) based on the scree plot, the additional proportion of variance explained by each factor (>0.10), and the independence of resultant factors. Day and night light variables were calculated by averaging light values across the respective time bins. Internal consistency of both day (Cronbach’s α = 0.98) and night (α = 0.93) light variables was very good.

Reliability of day and night light measurements was assessed in a subset of the actigraphy sample (*n* = 2,988) that completed a series of four repeated actigraphy assessments. These repeated assessments occurred on average 3.23 years (SD = 0.65) after the main assessment and took place over the course of a year with three months between each measurement. A linear mixed-effects model with a crossed random effect structure including participant, assessment (one to four), and time of day (day and night) was used to estimate the intra-class correlation (ICC) of (log-transformed) light. Due to the large positive skew in both day and night light variables (skewness > 1 for both), both variables were converted into categorical predictors for analysis by dividing them into four equal-sized quartiles (*n*_Q1-Q4_ = 21,693) in ascending brightness.

**Outcome variable definitions**

Psychiatric case/control outcomes and continuous symptom severity scores were defined based on participant responses to the UK Biobank Mental Health Questionnaire (MHQ) following guidelines established by Davis, Coleman ^5^.

***Lifetime major depressive disorder***

Lifetime major depressive disorder cases were defined according to the Composite International Diagnostic Interview (CIDI) lifetime depression module based on the DSM definition of major depressive disorder as well as professional diagnosis of major depression^6^.

*Cases*

- Responded ‘yes’ to either or both core symptom questions:
  - “Have you ever had a time in your life when you felt sad, blue, or depressed for two weeks or more in a row?? (data-field 20446)
  - “Have you ever had a time in your life lasting two weeks or more when you lost interest in most things like hobbies, work, or activities that usually give you pleasure?” (data-field 20441).
- AND responded “most of the day” or “all day long” to the question: “How much of the day did these feelings usually last?” (data-field 20436)
- AND responded “almost every day” or “every day” to the question: “Did you feel this way?” (data-field 20439)
- AND responded “somewhat” or “a lot” to the question: "Think about your roles at the time of this episode, including study / employment, childcare and housework, leisure pursuits. How much did these problems interfere with your life or activities?" (data-field 20440)
- AND endorsed ≥ 5 symptoms:
  - “Have you ever had a time in your life when you felt sad, blue, or depressed for two weeks or more in a row?” (data-field 20446)
  - “Have you ever had a time in your life lasting two weeks or more when you lost interest in most things like hobbies, work, or activities that usually give you pleasure?” (data-field 20441)
  - “Did you feel more tired out or low on energy than is usual for you?” (data-field 20449)
  - “Did you gain or lose weight without trying, or did you stay about the same weight?” (data-field 20536). Responding “gained”, “lost” or “both gained and lost” counted as endorsement.
  - “Did your sleep change?” (data-field 20532)
  - “Did you have a lot more trouble concentrating than usual?” (data-field 20435)
  - “People sometimes feel down on themselves, no good, worthless. Did you feel this way?” (data-field 20450)
  - “Did you think a lot about death - either your own, someone else's or death in general?” (data-field 20437)
- OR reported a professional diagnosis of major depression (data-field 20544 = 11)

*Controls*

- Responded “no” to both core symptom questions:
  - “Have you ever had a time in your life when you felt sad, blue, or depressed for two weeks or more in a row?” (data-field 20446)
  - “Have you ever had a time in your life lasting two weeks or more when you lost interest in most things like hobbies, work, or activities that usually give you pleasure?” (data-field 20441).
- Did not report a professional diagnosis of depression (data-field 20544)
- Had a PHQ-9 score of ≤ 5

***Lifetime generalized anxiety disorder***

Lifetime generalized anxiety disorder cases according to the CIDI lifetime GAD module based on the DSM-IV definition of GAD^6^.

*Cases*

- Responded yes to the question: “Have you ever had a period lasting one month or longer when most of the time you felt worried, tense, or anxious?” (data-field 20421)
- AND responded ≥ 6 months or “all my life/as long as I can remember” to the question: “What is the longest period of time that this kind of worrying has ever continued?” (data-field 20420)
- AND responded “yes” to the question: “Please think of the period in your life when you have felt worried, tense, anxious, or more worried than most people would in your situation. This could be in the past, or it could be continuing now. Did you worry most days?” (data-field 20538)
- AND responded
  - “more than most” to the question: “People differ a lot in how much they worry about things. Did you ever have a time when you worried a lot more than most people would in your situation?” (data-field 20425)
  - OR “stronger than most” to the question: “Please think of the period in your life when you have felt worried, tense, anxious, or more worried than most people would in your situation. This could be in the past, or it could be continuing now. During that period, was your worry stronger than in other people?” (data-field 20542)
- AND responded
  - “more than one thing” to the question: “Please think of the period in your life when you have felt worried, tense, anxious, or more worried than most people would in your situation. This could be in the past, or it could be continuing now. Did you usually worry about one particular thing, such as your job security or the failing health of a loved one, or more than one thing?” (data-field 20543)
  - OR “yes” to the question: “Please think of the period in your life when you have felt worried, tense, anxious, or more worried than most people would in your situation. This could be in the past, or it could be continuing now. Did you ever have different worries on your mind at the same time?” (data-field 20540)
- AND responded:
  - “yes” to the question: “Please think of the period in your life when you have felt worried, tense, anxious, or more worried than most people would in your situation. This could be in the past, or it could be continuing now. Did you find it difficult to stop worrying?” (data-field 20541)
  - OR “sometimes“ or “often” to the question: “Please think of the period in your life when you have felt worried, tense, anxious, or more worried than most people would in your situation. This could be in the past, or it could be continuing now. How often was your worry so strong that you couldn't put it out of your mind no matter how hard you tried?” (data-field 20539)
  - OR “sometimes” or “often” to the question: “Please think of the period in your life when you have felt worried, tense, anxious, or more worried than most people would in your situation. This could be in the past, or it could be continuing now. How often did you find it difficult to control your worry?” (data-field 20537)
- AND responded “somewhat” or “a lot” to the question: “Think about your roles at the time of this episode, including study / employment, childcare and housework, leisure pursuits. How much did these problems interfere with your life or activities?” (data-field 20418)
- AND endorsed three or more somatic symptoms by responding “yes” to the questions: “When you were worried or anxious, were you also:”
  - “Restless?” (data-field 20426)
  - “Keyed up or on edge?” (data-field 20423)
  - “Easily tired?” (data-field 20429)
  - “Having difficulty keeping on your mind what you were doing?” (data-field 20419)
  - “More irritable than usual?” (data-field 20422)
  - “Having tense, sore or aching muscles?” (data-field 20417)
  - “Often having trouble falling or staying asleep?” (data-field 20427)

*Controls*

- Did not meet the above criteria
- GAD-7 score of < 5

***Lifetime bipolar disorder***

Lifetime bipolar disorder cases were defined according to DSM-IV guidelines^7,8^.

*Cases*

- Responded “yes” to either or both core symptom questions:
  - “Have you ever had a period of time when you were feeling so good, "high", "excited", or "hyper" that other people thought you were not your normal self or you were so "hyper" that you got into trouble?” (data-field 20501)
  - “Have you ever had a period of time when you were so irritable that you found yourself shouting at people or starting fights or arguments?” (data-field 20502)
- AND endorsed four or more features of bipolar disorder from:
  - “I was more active than usual” (data-field 20548(1))
  - “I was more talkative than usual” (data-field 20548(2))
  - “I needed less sleep than usual” (data-field 20548(3))
  - “I was more creative or had more ideas than usual” (data-field 20548(4))
  - “I was more restless than usual” (data-field 20548(5))
  - I was more confident than usual (data-field 20548(6))
  - “My thoughts were racing” (data-field 20548(7))
  - “I was easily distracted” (data-field 20548(8))
  - “Have you ever had a period of time when you were feeling so good, "high", "excited", or "hyper" that other people thought you were not your normal self or you were so "hyper" that you got into trouble?” (data-field 20501)
- AND responded “a week or more” when asked: “What is the longest time that these "high" or "irritable" periods have lasted?” (data-field 20492)
- AND responded “needed treatment or caused problems with work, relationships, finances, the low or other aspects of life” to the question: “How much of a problem have these "high" or "irritable" periods caused you?” (data-field 20493)
- OR reported a professional diagnosis of bipolar disorder (data-field 20544 = 10)

*Controls*

- Did not meet the above criteria
- Did not report a professional diagnosis of bipolar disorder (data-field 20544 = 10)

***Lifetime psychosis***

Lifetime psychosis cases were defined using participant responses to the MHQ items which were adapted from the CIDI^9^.

*Cases*

- Responded “yes” to any of the following questions:
  - “Did you ever believe that a strange force was trying to communicate directly with you by sending special signs or signals that you could understand but that no one else could understand (for example through the radio or television)?” (data-field 20474)
  - “Did you ever believe that there was an unjust plot going on to harm you or to have people follow you, and which your family and friends did not believe existed?” (data-field 20468)
  - “Did you ever see something that wasn't really there that other people could not see?” (data-field 20471)
  - “Did you ever hear things that other people said did not exist, like strange voices coming from inside your head talking to you or about you, or voices coming out of the air when there was no one around?” (data-field 20463)
- OR reported a professional diagnosis of schizophrenia (data-field 20544 = 2) or psychotic illness (data-field 20544 = 3)

*Controls*

- Did not endorse any of the four psychotic experiences
- Did not report a professional diagnosis of schizophrenia (data-field 20544 = 2) or psychotic illness (data-field 20544 = 3)

***Post-traumatic stress disorder***

PTSD cases and controls were defined using the PTSD-Checklist-6 (PCL-6) with a clinical case-control threshold of 14 that is validated against the CIDI^10^. See below (appendix X p Y *** for PCL-6 score definition)

*Cases*

- Responded “a little bit”, “moderately”, “quite a bit” or “extremely” to any of the following questions: “Next is a list of problems and complaints that people sometimes have in response to such extremely stressful experiences. Please indicate how much you have been bothered by that problem in the past month:”
  - “Repeated, disturbing memories, thoughts or images of a stressful experience?” (data-field 20497)
  - “Feeling very upset when something reminded you of a stressful experience?” (data-field 20498)
  - “Avoiding activities or situations because they reminded you of a stressful experience?” (data-field 20495)
- AND total PCL-6 symptom severity score ≥ 14

*Controls*

- Responded to data-fields 20487, 20498 and 20495
- AND total PCL-6 symptom severity score < 14

***Self-harm***

Self-harm cases and controls were defined by “yes” or “no” answers to the following question: “Have you ever deliberately harmed yourself, whether or not you meant to end your life?” (data-field 20480)

***PHQ-9 Score***

The Patient Health Questionnaire-9 (PHQ-9) scale is a measure of depressive symptom severity^11^. A total score (range 0-27) was calculated by summing participant responses to the following nine items: “Over the last two weeks, how often have you been bothered by any of the following problems?”:

- “Little interest or pleasure in doing things (data-field 20514)
- “Feeling down depressed or hopeless” (data-field 20510)
- “Trouble falling asleep or sleeping too much” (data-field 20517)
- “Poor appetite or overeating” (data-field 20511)
- “Feeling bad about yourself or that you are a failure or have let yourself or your family down” (data-field 20507)
- “Trouble concentrating on things, such as reading the newspaper or watching television” (data-field 20508)
- “Moving or speaking so slowly that other people could have noticed? Or the opposite – being so fidgety or restless that you have been moving around a lot more than usual” (data-field 20518)
- “Thoughts that you would be better off dead or of hurting yourself in some way” (data-field 20513)

Response options were “not at all” (coded as 0), “several days” (1), “more than half the days” (2) and “nearly every day” (3).

***GAD-7 Score***

The Generalized Anxiety Disorder-7 (GAD-7) scale is a measure of anxiety symptom severity^11,12^. A total score (range 0-21) was calculated by summing participant responses to the following seven items: “Over the last two weeks, how often have you been bothered by any of the following problems?”:

- “Feeling nervous, anxious or on edge” (data-field 20506)
- “Not being able to stop or control worrying” (data-field 20509)
- “Worrying too much about different things” (data-field 20520)
- “Trouble relaxing” (data-field 20515)
- “Being so restless that it is hard to sit still” (data-field 20516)
- “Becoming so easily annoyed or irritable” (data-field 20505)
- “Feeling afraid as if something awful might happen” (data-field 20512)

Response options were “not at all” (coded as 0), “several days” (1), “more than half the days” (2) and “nearly every day” (3).

***PCL-6 Score***

The PTSD-Checklist-6 (PCL-6) scale is a measure of PTSD symptom severity^10^. A total score (range 6-29) was calculated by summing participant responses to the following six items: “Please indicate how much you have been bothered by that problem in the past month:”

- “Repeated, disturbing memories, thoughts or images of a stressful experience?” (data-field 20497)
- “Feeling very upset when something reminded you of a stressful experience?” (data-field 20498)
- “Avoiding activities or situations because they reminded you of a stressful experience?” (data-field 20495)
- “Feeling distant or cut off from other people?” (data-field 20496)
- “Feeling irritable or having angry outbursts?” (data-field 20494)
- “Trouble concentrating on things, such as reading the newspaper or watching television” (data-field 20508)

Response options were “not at all” (coded as 1), “a little bit” (2), “moderately” (3), “quite a bit” (4) and “extremely” (5) except for data-field 20508 which was coded: “not at all” (coded as 1), “several days” (2), “more than half the days” (3) and “nearly every day” (4).

***Wellbeing score***

A total wellbeing score (range 3-17) was calculated by summing participant responses to the following three items:

- “In general how happy are you?” (data-field 20458)
- “In general how happy are you with your health?” (data-field 20459)
- “To what extent do you feel your life to be meaningful?” (data-field 20460)

Response options for data-fields 20458 and 20459 were from “extremely unhappy” (coded as 1) to “extremely happy” (6). Response options for data-field 20460 were from “not at all” (1) to “an extreme amount” (5).

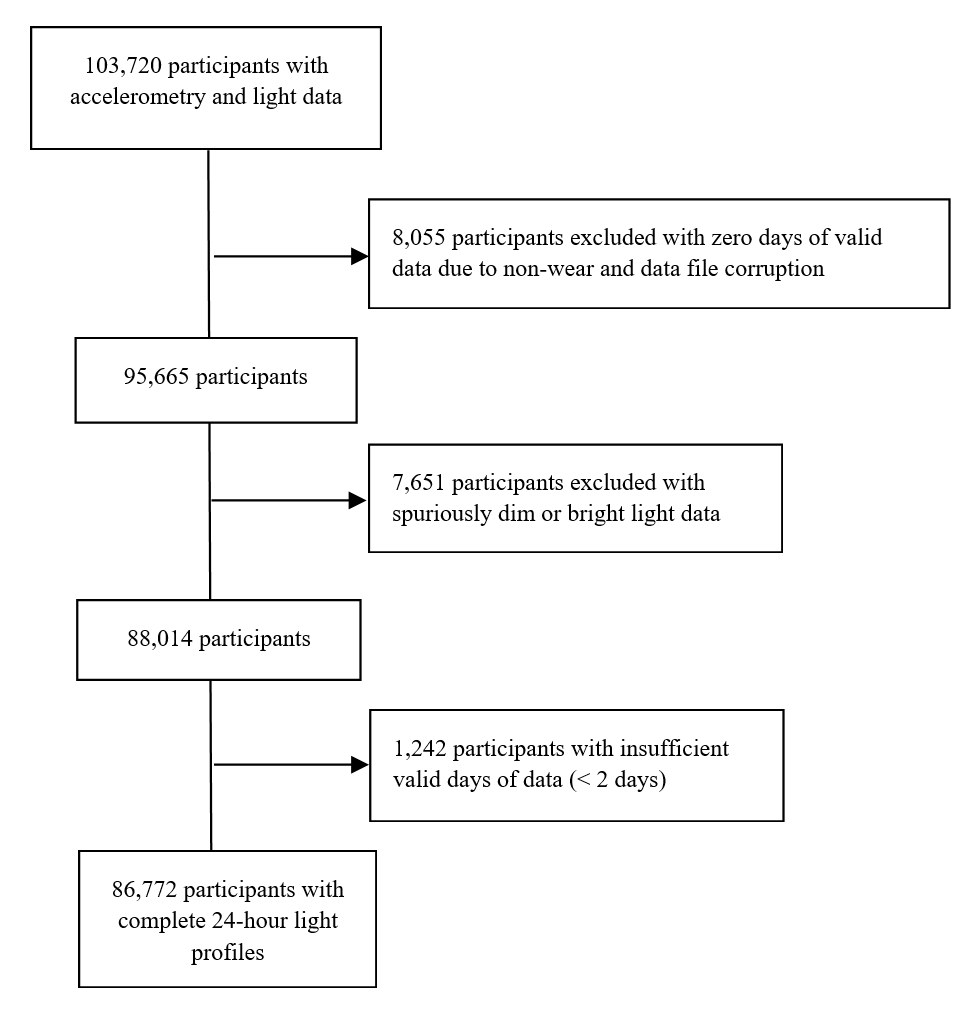

**Supplementary Figure 1.** Flow diagram of light data study sample

**Supplementary Table 1.** Summary statistics of psychiatric outcomes among the light data sample

| ***n* = 86,772** | ***Major depressive disorder*** | **Self-harm** | **Generalized anxiety disorder** | **PTSD** | **Bipolar disorder** | **Psychosis** |
| --- | --- | --- | --- | --- | --- | --- |
| **Control *n* (%)** | 25,509 (29·4%) | 58,670 (67·6%) | 46,554 (53·7%) | 57,907 (66·7%) | 45,197 (52·1%) | 57,785 (66·6%) |
| **Case *n* (%)** | 18,933 (21·8%) | 2,710 (3·1%) | 3,279 (3·8%) | 3,535 (4·1%) | 1,192 (1·4%) | 3,110 (3·6%) |
| **Missing *n* (%)** | 42,330 (48·8%) | 25,392 (29·3%) | 36,939 (42·6%) | 25,330 (29·2%) | 40,383 (46·5%) | 25,877 (29·8%) |

Missing data include those in the light data sample who did not complete the MHQ, answer relevant questions for case/control definitions or meet case/control criteria (see eAppendix).

**Supplementary Table 2.** Summary statistics of continuous symptom severity scales and wellbeing among the light data sample

| ***n* = 86,772** | ***PHQ-9*** | **GAD-7** | **PCL-6** | **Wellbeing** |
| --- | --- | --- | --- | --- |
| **Mean (SD)** | 2·64 (3·54) | 2·07 (3·31) | 10·1 (3·29) | 8·30 (1·26) |
| **Median [Q1, Q3]** | 2 [0, 4] | 0 [0, 3] | 9 [8, 11] | 8 [8, 9] |
| **Missing n (%)** | 26,216 (30·2%) | 25,986 (29·9%) | 59,816 (68·9%) | 26,868 (31·0%) |
| ***n* (%)** | 60,556 (69·8%) | 60,786 (70·1%) | 26,956 (31·1%) | 59,904 (69·0%) |

Missing data include those in the light data sample who did not complete the MHQ or answer relevant questions for continuous symptom severity scale definitions (see eAppendix).

**Supplementary Table 3. Associations between night light exposure and symptom severity scales**

|  | **Model 1**  **Beta (95% CI)** | | | | **Model 2**  **aBeta (95% CI)*** | | | | **Model 3**  **aBeta (95% CI)^†^** | | | |
| --- | --- | --- | --- | --- | --- | --- | --- | --- | --- | --- | --- | --- |
| **Night light** | **2^nd^**  **Quartile** | **3^rd^  Quartile** | **4^th^ Quartile** | χ^2^ ***p*** | **2^nd^ Quartile** | **3^rd^ Quartile** | **4^th^ Quartile** | χ^2^ ***p*** | **2^nd^**  **Quartile** | **3^rd^**  **Quartile** | **4^th^**  **Quartile** | χ^2^ ***p*** |
| **PHQ-9** | 0·02 (-0·00 – 0·04) | **0**·**08 (0**·**06 – 0**·**10)** | **0**·**13 (0**·**11 – 0**·**16)** | **<0**·**0001** | 0·01 (-0·01 – 0·03) | **0**·**06 (0**·**04 – 0**·**08)** | **0**·**13 (0**·**11 – 0**·**16)** | **<0**·**0001** | 0·01 (-0·01 – 0·03) | **0**·**07 (0**·**05 – 0**·**09)** | **0**·**13 (0**·**11 – 0**·**15)** | **<0**·**0001** |
| **Wellbeing** | **-0**·**02 (-0**·**05 – -0**·**01)** | **-0**·**07 (-0**·**10 – -0**·**05)** | **-0**·**11 (-0**·**14 – -0**·**09)** | **<0**·**0001** | -0·02 (-0·04 – 0·00) | **-0**·**06 (-0**·**09 – -0**·**04)** | **-0**·**11 (-0**·**14 – -0**·**09)** | **<0**·**0001** | -0·02 (-0·04 – 0·00) | **-0**·**07 (-0**·**09 – -0**·**05)** | **-0**·**11 (-0**·**13 – -0**·**08)** | **<0**·**0001** |
| **PCL-6** | 0·03 (-0·01 – 0·06) | **0**·**08 (0**·**04 – 0**·**11)** | **0**·**12 (0**·**08 – 0**·**15)** | **<0**·**0001** | 0·02 (-0·01 – 0·05) | **0**·**06 (0**·**03 – 0**·**10)** | **0**·**11 (0**·**08 – 0**·**15)** | **<0**·**0001** | 0·02 (-0·02 – 0·05) | **0**·**06 (0**·**03 – 0**·**10)** | **0**·**11 (0**·**08 – 0**·**14)** | **<0**·**0001** |
| **GAD-7** | 0·02 (-0·00 – 0·04) | **0**·**05 (0**·**03 – 0**·**07)** | **0**·**07 (0**·**05 – 0**·**09)** | **<0**·**0001** | 0·01 (-0·01 – 0·03) | **0**·**04 (0**·**02 – 0**·**06)** | **0**·**07 (0**·**05 – 0**·**09)** | **<0**·**0001** | 0·01 (-0·01 – 0·03) | **0**·**04 (0**·**02 – 0**·**06)** | **0**·**07 (0**·**05 – 0**·**09)** | **<0**·**0001** |

Note: linear regression was used for all outcomes and standardized betas are presented with their 95% confidence intervals for each ascending night light quartile relative to the low light (Q1) referent. Model 1 is unadjusted. *Model 2 adjusted for age, sex, ethnicity, and photoperiod as a measure of seasonality. **^†^**Model 3 is additionally adjusted for employment and physical activity. Bolded coefficients indicate significance at the .05 level.

**Supplementary Table 4. Associations between day light exposure and symptom severity scales**

|  | **Model 1**  **Beta (95% CI)** | | | | **Model 2**  **aBeta (95% CI)*** | | | | **Model 3**  **aBeta (95% CI)^†^** | | | |
| --- | --- | --- | --- | --- | --- | --- | --- | --- | --- | --- | --- | --- |
| **Night light** | **2^nd^**  **Quartile** | **3^rd^  Quartile** | **4^th^ Quartile** | χ^2^ ***p*** | **2^nd^ Quartile** | **3^rd^ Quartile** | **4^th^ Quartile** | χ^2^ ***p*** | **2^nd^**  **Quartile** | **3^rd^**  **Quartile** | **4^th^**  **Quartile** | χ^2^ ***p*** |
| **PHQ-9** | -0·02 (-0·04 – 0·01) | **-0**·**03 (-0**·**05 – -0**·**00)** | **-0**·**11 (-0**·**13 – -0**·**08)** | **<0**·**0001** | **-0**·**03 (-0**·**05 – -0**·**01)** | **-0**·**06 (-0**·**08 – -0**·**04)** | **-0**·**14 (-0**·**17 – -0**·**12)** | **<0**·**0001** | -0·02 (-0·04 – 0·00) | **-0**·**04 (-0**·**06 – -0**·**01)** | **-0**·**09 (-0**·**12 – -0**·**07)** | **<0**·**0001** |
| **Wellbeing** | 0·01 (-0·01 – 0·04) | **0**·**03 (0**·**01 – 0**·**06)** | **0**·**09 (0**·**07 – 0**·**12)** | **<0**·**0001** | **0**·**03 (0**·**00 – 0**·**05)** | **0**·**07 (0**·**04 – 0**·**09)** | **0**·**14 (0**·**12 – 0**·**17)** | **<0**·**0001** | 0·01 (-0·01 – 0·03) | **0**·**04 (0**·**01 – 0**·**06)** | **0**·**08 (0**·**05 – 0**·**11)** | **<0**·**0001** |
| **PCL-6** | -0·03 (-0·06 – 0·01) | -0·03 (-0·07 – 0·00) | **-0**·**07 (-0**·**11 – -0**·**04)** | **0**·**0004** | **-0**·**05 (-0**·**08 – -0**·**01)** | **-0·06 (-0·10 – -0·03)** | **-0**·**11 (-0**·**15 – -0**·**07)** | **<0**·**0001** | **-0**·**03 (-0**·**07 – -0**·**00)** | **-0**·**05 (-0**·**08 – -0**·**01)** | **-0**·**08 (-0**·**12 – -0**·**04)** | **0**·**002** |
| **GAD-7** | -0·01 (-0·03 – 0·02) | 0·01 (-0·02 – 0·03) | **-0**·**06 (-0**·**08 – -0**·**04)** | **<0**·**0001** | -0·02 (-0·04 – 0·01) | -0·01 (-0·03 – 0·01) | **-0**·**07 (-0**·**10 – -0**·**04)** | **<0**·**0001** | -0·01 (-0·03 – 0·01) | -0·00 (-0·03 – 0·02) | **-0**·**05 (-0**·**08 – -0**·**03)** | **<0**·**0001** |

Note: linear regression was used for all outcomes and standardized betas are presented with their 95% confidence intervals for each ascending day light quartile relative to the low light (Q1) referent. Model 1 is unadjusted. *Model 2 adjusted for age, sex, ethnicity, and photoperiod as a measure of seasonality. **^†^**Model 3 is additionally adjusted for employment and physical activity. Bolded coefficients indicate significance at the .05 level.

**Supplementary Table 5.** Associations of day and night light exposure with psychiatric outcomes excluding shift workers

|  | **Night light**  **aOR (95% CI)*** | | | | **Day light**  **aOR (95% CI)*** | | | |
| --- | --- | --- | --- | --- | --- | --- | --- | --- |
| **Outcome** | **2^nd^**  **Quartile** | **3^rd^  Quartile** | **4^th^ Quartile** | χ^2^ ***p*** | **2^nd^ Quartile** | **3^rd^ Quartile** | **4^th^ Quartile** | χ^2^ ***p*** |
| **Major depressive disorder** | 1·05  (0·99 – 1·11) | **1·18**  **(1·12 – 1·26)** | **1·30**  **(1·23 – 1·38)** | **<0·0001** | **0·93**  **(0·88 – 0·99)** | **0·88**  **(0·83 – 0·94)** | **0·82**  **(0·76 – 0·88)** | **<0·0001** |
| **Self-harm** | 1·02  (0·90 – 1·14) | 1·01  (0·90 – 1·14) | **1·28**  **(1·14 – 1·44)** | **<0·0001** | **0·83**  **(0·74 – 0·93)** | **0·78**  **(0·69 – 0·88)** | **0·75**  **(0·65 – 0·86)** | **<0·0001** |
| **Generalized anxiety disorder** | 1·08  (0·97 – 1·20) | **1·20**  **(1·08 – 1·34)** | **1·27**  **(1·14 – 1·41)** | **<0·0001** | 1·00  (0·90 – 1·11) | 0·93  (0·83 – 1·05) | 0·93  (0·82 – 1·06) | 0·49 |
| **PTSD** | 1·00  (0·90 – 1·11) | **1·14**  **(1·03 – 1·27)** | **1·31**  **(1·18 – 1·45)** | **<0·0001** | 0·92  (0·83 – 1·02) | **0·88**  **(0·79 – 0·98)** | **0·82**  **(0·73 – 0·93)** | **0·02** |
| **Bipolar disorder** | 1·10  (0·93 – 1·31) | 1·09  (0·91 – 1·30) | **1·20**  **(1·01 – 1·43)** | 0·25 | 0·90  (0·76 – 1·08) | 0·99  (0·82 – 1·18) | 0·87  (0·70 – 1·07) | 0·38 |
| **Psychosis** | 1·02  (0·92 – 1·14) | 1·09  (0·98 – 1·22) | **1·19**  **(1·06 – 1·32)** | **0·009** | 0·98  (0·88 – 1·09) | **0·83**  **(0·74 – 0·93)** | **0·71**  **(0·62 – 0·81)** | **<0·0001** |

Note: logistic regression was used for all outcomes and odds ratios (ORs) are presented with their 95% confidence intervals for each ascending night light quartile relative to the low light (Q1) referent. *Statistical model used the same covariates as Model 3, adjusting for age, sex, ethnicity, photoperiod as a measure of seasonality, employment, and physical activity and excluded participants engaged in shift work. Bolded coefficients indicate significance at the .05 level.

**Supplementary Table 6.** Associations of day and night light exposure with symptom severity scales excluding shift workers

|  | **Night light**  **aBeta (95% CI)*** | | | | **Day light**  **aBeta (95% CI)*** | | | |
| --- | --- | --- | --- | --- | --- | --- | --- | --- |
| **Outcome** | **2^nd^**  **Quartile** | **3^rd^  Quartile** | **4^th^ Quartile** | χ^2^ ***p*** | **2^nd^ Quartile** | **3^rd^ Quartile** | **4^th^ Quartile** | χ^2^ ***p*** |
| **PHQ-9** | 0·01  (-0·01 – 0·04) | **0·07**  **(0·05 – 0·09)** | **0·13**  **(0·11 – 0·15)** | **<0·0001** | -0·02  (-0·04 – 0·01) | **-0·04**  **(-0·06 – -0·01)** | **-0·10**  **(-0·12 – -0·07)** | **<0·0001** |
| **Wellbeing** | -0·02  (-0·04 – 0·01) | **-0·07**  **(-0·09 – -0·05)** | **-0·10**  **(-0·12 – -0·08)** | **<0·0001** | 0·01  (-0·01 – 0·03) | **0·03**  **(0·01 – 0·06)** | **0·07**  **(0·05 – 0·10)** | **<0·0001** |
| **PCL-6** | 0·01  (-0·02 – 0·05) | **0·06**  **(0·03 – 0·10)** | **0·10**  **(0·07 – 0·14)** | **<0·0001** | **-0·04**  **(-0·07 – -0·00)** | **-0·05**  **(-0·09 – -0·02)** | **-0·08**  **(-0·12 – -0·04)** | **0·002** |
| **GAD-7** | 0·01  (-0·01 – 0·03) | **0·04**  **(0·02 – 0·06)** | **0·07**  **(0·05 – 0·09)** | **<0·0001** | -0·00  (-0·03 – 0·02) | 0·00  (-0·02 – 0·03) | **-0·05**  **(-0·08 – -0·02)** | **<0·0001** |

Note: linear regression was used for all outcomes and standardized betas are presented with their 95% confidence intervals for each ascending night light quartile relative to the low light (Q1) referent. *Statistical model used the same covariates as Model 3, adjusting for age, sex, ethnicity, photoperiod as a measure of seasonality, employment, and physical activity and excluded participants engaged in shift work. Bolded coefficients indicate significance at the .05 level.

**Supplementary Table 7.** Associations of day and night light exposure with symptom severity scales excluding participants meeting disorder criteria

|  | **Night light**  **aBeta (95% CI)*** | | | | **Day light**  **aBeta (95% CI)*** | | | |
| --- | --- | --- | --- | --- | --- | --- | --- | --- |
| **Outcome†** | **2^nd^**  **Quartile** | **3^rd^  Quartile** | **4^th^ Quartile** | χ^2^ ***p*** | **2^nd^ Quartile** | **3^rd^ Quartile** | **4^th^ Quartile** | χ^2^ ***p*** |
| **PHQ-9** | 0·01 (-0·02 – 0·03) | **0·05 (0·03 – 0·08)** | **0·11 (0·08 – 0·13)** | **<0·0001** | -0·03 (-0·06 – -0·00) | **-0·04 (-0·07 – -0·02)** | **-0·10 (-0·14 – -0·07)** | **<0·0001** |
| **Wellbeing** | -0·02 (-0·04 – 0·00) | **-0·07 (-0·09 – -0·05)** | **-0·11 (-0·13 – -0·08)** | **<0·0001** | 0·01 (-0·02 – 0·03) | **0·04 (0·01 – 0·06)** | **0·08 (0·05 – 0·11)** | **<0·0001** |
| **PCL-6** | **0·04 (0·00 – 0·07)** | **0·06 (0·02 – 0·09)** | **0·10 (0·06 – 0·13)** | **<0·0001** | -0·02 (-0·06 – 0·02) | **-0·05 (-0·09 – -0·01)** | **-0·08 (-0·12 – -0·03)** | **0·006** |
| **GAD-7** | 0·01 (-0·01 – 0·03) | **0·03 (0·01 – 0·05)** | **0·06 (0·03 – 0·08)** | **<0·0001** | -0·01 (-0·04 – 0·01) | 0·00 (-0·02 – 0·03) | **-0·05 (-0·08 – -0·02)** | **<0·0001** |

Note: linear regression was used for all outcomes and standardized betas are presented with their 95% confidence intervals for each ascending night light quartile relative to the low light (Q1) referent. *Statistical model used the same covariates as Model 3, adjusting for age, sex, ethnicity, photoperiod as a measure of seasonality, employment, and physical activity. **^†^**PHQ-9 model excluded participants meeting MDD criteria; PCL-6 model excluded participants meeting PTSD criteria; GAD-7 model excluded participants meeting GAD criteria; and wellbeing model excluded participants meeting MDD, PTSD, GAD, bipolar disorder, or psychosis criteria. Bolded coefficients indicate significance at the .05 level.

**Supplementary Table 8.** Associations of day and night light exposure with psychiatric outcomes adjusting for sleep variables

|  | **Night light**  **aOR (95% CI)*** | | | | **Day light**  **aOR (95% CI)*** | | | |
| --- | --- | --- | --- | --- | --- | --- | --- | --- |
| **Outcome** | **2^nd^**  **Quartile** | **3^rd^  Quartile** | **4^th^ Quartile** | χ^2^ ***p*** | **2^nd^ Quartile** | **3^rd^ Quartile** | **4^th^ Quartile** | χ^2^ ***p*** |
| **Major depressive disorder** | 1·04 (0·98 – 1·09) | **1**·**17 (1**·**11 – 1**·**24)** | **1**·**28 (1**·**21 – 1**·**36)** | **<0**·**0001** | **0·93 (0·88 – 0·99)** | **0**·**88 (0**·**83 – 0**·**94)** | **0·82 (0**·**76 – 0**·**87)** | **<0**·**0001** |
| **Self-harm** | 0·98 (0·88 – 1·10) | 0·97 (0·87 – 1·09) | **1**·**19 (1**·**06 – 1**·**33)** | **0**·**0008** | **0·86 (0·77 – 0·96)** | **0·80 (0·71 – 0·90)** | **0**·**79 (0**·**69 – 0**·**90)** | **0·0006** |
| **Generalized anxiety disorder** | 1·07  (0·96 – 1·18) | **1**·**18**  **(1**·**07 – 1**·**31)** | **1**·**18**  **(1**·**06 – 1**·**31)** | **0·003** | 1·00  (0·90 – 1·10) | 0·94  (0·84 – 1·05) | 0·98  (0·86 – 1·10) | 0·70 |
| **PTSD** | 1·00  (0·90 – 1·11) | **1**·**14**  **(1**·**03 – 1**·**26)** | **1**·**26**  **(1**·**14 – 1**·**40)** | **<0**·**0001** | 0·94  (0·85 – 1·04) | 0·91  (0·82 – 1·01) | **0**·**84**  **(0**·**75 – 0**·**95)** | **0·04** |
| **Bipolar disorder** | 1·10  (0·93 – 1·30) | 1·11  (0·94 – 1·31) | 1·16  (0·98 – 1·38) | 0·37 | 0·88  (0·74 – 1·03) | 0·97  (0·82 – 1·16) | 0·87  (0·71 – 1·06) | 0·27 |
| **Psychosis** | 1·03  (0·93 – 1·15) | 1·11  (1·00 – 1·23) | **1**·**12**  **(1**·**01 – 1**·**25)** | 0·09 | 0·98  (0·89 – 1·09) | **0**·**83**  **(0**·**74 – 0**·**93)** | **0**·**72**  **(0**·**63 – 0**·**81)** | **<0**·**0001** |

Note: logistic regression was used for all outcomes and odds ratios (ORs) are presented with their 95% confidence intervals for each ascending night light quartile relative to the low light (Q1) referent. *Model 4 covariates adjusted for sleep duration and sleep efficiency as well as the covariates in Model 3: age, sex, ethnicity, photoperiod as a measure of seasonality, employment, and physical activity. Bolded coefficients indicate significance at the .05 level.

**Supplementary Table 9.** Associations of day and night light with symptom severity scales adjusting for sleep variables

|  | **Night light**  **aBeta (95% CI)*** | | | | **Day light**  **aBeta (95% CI)*** | | | |
| --- | --- | --- | --- | --- | --- | --- | --- | --- |
| **Outcome** | **2^nd^**  **Quartile** | **3^rd^  Quartile** | **4^th^ Quartile** | χ^2^ ***p*** | **2^nd^ Quartile** | **3^rd^ Quartile** | **4^th^ Quartile** | χ^2^ ***p*** |
| **PHQ-9** | 0·01  (-0·01 – 0·03) | **0·06**  **(0·03 – 0·08)** | **0·10**  **(0·07 – 0·12)** | **<0·0001** | -0·01  (-0·03 – 0·01) | **-0·03**  **(-0·05 – -0·00)** | **-0·08**  **(-0·11 – -0·05)** | **<0·0001** |
| **Wellbeing** | -0·02  (-0·04 – 0·00) | **-0·06**  **(-0·08 – -0·04)** | **-0·07**  **(-0·10 – -0·05)** | **<0·0001** | 0·00  (-0·02 – 0·03) | **0·03**  **(0·00 – 0·05)** | **0·06**  **(0·04 – 0·09)** | **<0·0001** |
| **PCL-6** | 0·01  (-0·02 – 0·05) | **0·05**  **(0·02 – 0·09)** | **0·09**  **(0·05 – 0·12)** | **<0·0001** | -0·03  (-0·06 – 0·00) | **-0·04**  **(-0·08 – -0·01)** | **-0·07**  **(-0·11 – -0·03)** | **0·008** |
| **GAD-7** | 0·01  (-0·01 – 0·03) | **0·03**  **(0·01 – 0·06)** | **0·06**  **(0·03 – 0·08)** | **<0·0001** | -0·01  (-0·03 – 0·01) | 0·00  (-0·02 – 0·03) | **-0·05**  **(-0·08 – -0·02)** | **0·0001** |

Note: linear regression was used for all outcomes and standardized betas are presented with their 95% confidence intervals for each ascending night light quartile relative to the low light (Q1) referent. *Model 4 covariates adjusted for sleep duration and sleep efficiency as well as the covariates in Model 3: age, sex, ethnicity, photoperiod as a measure of seasonality, employment, and physical activity. Bolded coefficients indicate significance at the .05 level.

**Supplementary Table 10.** Associations of day and night light exposure with psychiatric outcomes adjusting for urbanicity

|  | **Night light**  **aOR (95% CI)*** | | | | **Day light**  **aOR (95% CI)*** | | | |
| --- | --- | --- | --- | --- | --- | --- | --- | --- |
| **Outcome** | **2^nd^**  **Quartile** | **3^rd^  Quartile** | **4^th^ Quartile** | χ^2^ ***p*** | **2^nd^ Quartile** | **3^rd^ Quartile** | **4^th^ Quartile** | χ^2^ ***p*** |
| **Major depressive disorder** | 1·04 (0·98 – 1·10) | **1**·**18 (1**·**12 – 1**·**25)** | **1**·**30 (1**·**23 – 1**·**38)** | **<0**·**0001** | **0·94 (0·89 – 0·99)** | **0**·**89 (0**·**84 – 0**·**94)** | **0·82 (0**·**77 – 0**·**88)** | **<0**·**0001** |
| **Self-harm** | 0·98 (0·88 – 1·10) | 1·00 (0·89 – 1·12) | **1**·**26 (1**·**13 – 1**·**41)** | **<0**·**0001** | **0·85 (0·76 – 0·94)** | **0·78 (0·69 – 0·87)** | **0**·**76 (0**·**66 – 0**·**87)** | **<0**·**0001** |
| **Generalized anxiety disorder** | 1·06  (0·96 – 1·18) | **1**·**20**  **(1**·**08 – 1**·**33)** | **1**·**22**  **(1**·**10 – 1**·**35)** | **0·0002** | 1·00  (0·90 – 1·11) | 0·94  (0·84 – 1·05) | 0·97  (0·86 – 1·10) | 0·65 |
| **PTSD** | 1·00  (0·91 – 1·11) | **1**·**16**  **(1**·**05 – 1**·**28)** | **1**·**34**  **(1**·**21 – 1**·**47)** | **<0**·**0001** | 0·94  (0·85 – 1·04) | 0·90  (0·81 – 1·00) | **0**·**83**  **(0**·**74 – 0**·**94)** | **0·03** |
| **Bipolar disorder** | 1·09  (0·93 – 1·29) | 1·12  (0·95 – 1·32) | **1·19**  **(1·01 – 1·41)** | 0·22 | 0·88  (0·74 – 1·03) | 0·96  (0·81 – 1·14) | 0·87  (0·71 – 1·06) | 0·32 |
| **Psychosis** | 1·03  (0·93 – 1·14) | **1·14**  **(1·03 – 1·27)** | **1**·**21**  **(1**·**09 – 1**·**35)** | **0·0006** | 0·96  (0·86 – 1·06) | **0**·**80**  **(0**·**72 – 0**·**90)** | **0**·**68**  **(0**·**60 – 0**·**77)** | **<0**·**0001** |

Note: logistic regression was used for all outcomes and odds ratios (ORs) are presented with their 95% confidence intervals for each ascending night light quartile relative to the low light (Q1) referent. *Model 5 covariates adjusted for urbanicity as well as the covariates in Model 3: age, sex, ethnicity, photoperiod as a measure of seasonality, employment, and physical activity. Bolded coefficients indicate significance at the .05 level.

**Supplementary Table 11.** Associations of day and night light exposure with symptom severity scales adjusting for urbanicity

|  | **Night light**  **aBeta (95% CI)*** | | | | **Day light**  **aBeta (95% CI)*** | | | |
| --- | --- | --- | --- | --- | --- | --- | --- | --- |
| **Outcome** | **2^nd^**  **Quartile** | **3^rd^  Quartile** | **4^th^ Quartile** | χ^2^ ***p*** | **2^nd^ Quartile** | **3^rd^ Quartile** | **4^th^ Quartile** | χ^2^ ***p*** |
| **PHQ-9** | 0·01  (-0·01 – 0·03) | **0·07**  **(0·05 – 0·09)** | **0·13**  **(0·11 – 0·15)** | **<0·0001** | -0·02  (-0·04 – 0·00) | **-0·04**  **(-0·06 – -0·01)** | **-0·09**  **(-0·12 – -0·07)** | **<0·0001** |
| **Wellbeing** | -0·02  (-0·04 – 0·00) | **-0·07**  **(-0·09 – -0·05)** | **-0·11**  **(-0·13 – -0·08)** | **<0·0001** | 0·01  (-0·01 – 0·03) | **0·03**  **(0·01 – 0·06)** | **0·08**  **(0·05 – 0·10)** | **<0·0001** |
| **PCL-6** | 0·02  (-0·02 – 0·05) | **0·06**  **(0·03 – 0·10)** | **0·11**  **(0·08 – 0·14)** | **<0·0001** | -0·03  (-0·07 – 0·00) | **-0·05**  **(-0·08 – -0·01)** | **-0·07**  **(-0·12 – -0·03)** | **0·005** |
| **GAD-7** | 0·01  (-0·01 – 0·03) | **0·04**  **(0·02 – 0·06)** | **0·07**  **(0·05 – 0·09)** | **<0·0001** | -0·01  (-0·03 – 0·01) | -0·00  (-0·02 – 0·02) | **-0·05**  **(-0·08 – -0·03)** | **<0·0001** |

Note: linear regression was used for all outcomes and standardized betas are presented with their 95% confidence intervals for each ascending night light quartile relative to the low light (Q1) referent. *Model 5 covariates adjusted for urbanicity as well as the covariates in Model 3: age, sex, ethnicity, photoperiod as a measure of seasonality, employment, and physical activity. Bolded coefficients indicate significance at the .05 level.

**References.**
